# Deep learning based treatment remission prediction to transcranial direct current stimulation in bipolar depression using EEG power spectral density

**DOI:** 10.1101/2024.07.16.24310445

**Authors:** Jijomon Chettuthara Moncy, Wenyi Xiao, Rachel D. Woodham, Ali-Reza Ghazi-Noori, Hakimeh Rezaei, Elvira Bramon, Philipp Ritter, Michael Bauer, Allan H. Young, Yong Fan, Cynthia H.Y. Fu

**Affiliations:** Department of Psychology, University of East London, London, UK; Technische Universität Dresden, Dresden, Germany; Department of Psychiatry, University College London, London, UK; Centre for Affective Disorders, Department of Psychological Medicine, Institute of Psychiatry, Psychology and Neuroscience, King’s College London, London, UK; National Institute for Health Research Biomedical Research Centre at South London and Maudsley NHS Foundation Trust, King’s College London, London, UK; South London and Maudsley NHS Foundation Trust, Bethlem Royal Hospital, Beckenham, UK; Department of Radiology, Perelman School of Medicine University of Pennsylvania Philadelphia, USA

**Keywords:** EEG, tDCS, power spectral density, deep learning, bipolar depression, predictors, prediction treatment remission

## Abstract

Bipolar disorder is characterized by marked changes in mood and activity levels and is a leading cause of disability worldwide. We sought to investigate the application of deep learning methods to electroencephalogram (EEG) signals to predict clinical remission after 6 weeks of home-based transcranial direct current stimulation (tDCS) treatment. Pre-treatment resting-state EEG acquired from 21 bipolar participants was used for this work. A hybrid 1DCNN and GRU model, with input from power spectral density values of theta, beta and gamma frequency bands of the AF7 and TP10 electrodes, achieved a treatment remission prediction accuracy of 78.5% (sensitivity 81.4%, specificity 74.64%).

## 1. Introduction

Bipolar disorder (BD) is characterised by marked changes in mood with depressive episodes, hypomanic or manic episodes, as well as potential mixed mood states, which are accompanied by alterations in sleep patterns, energy levels, and psychomotor activity [1]. BD consists of recurrent episodes, however, more time is spent in depressive episodes than in manic or hypomanic phases. Current treatments for BD have significant limitations, including variable response rates and potential side effects that can impact on long term adherence and clinical outcomes.

Transcranial direct current stimulation (tDCS) is a non-invasive neuromodulation technique that is a potential novel treatment for bipolar depression [2]. tDCS involves administering a low-amplitude direct current (0.5–2.0 mA) through surface scalp electrodes overlying targeted cortical areas. The applied current alters cortical excitability by adjusting the resting potential of neuronal membranes, in which the modulatory effects can persist beyond the duration of the electrical stimulation. The main side effects of tDCS are skin redness, tingling or itching sensations on the scalp, headache and drowsiness [3]. Our study follows a home-based protocol has demonstrated high efficacy, acceptability and safety for active relative to sham tDCS in unipolar depression [4]. Similarly, in bipolar depression, we found strong treatment effects and acceptability for a course of open-label active tDCS [5].

Identifying reliable biomarkers for clinical outcomes could enable more personalized and effective treatment approaches, reducing trial-and-error in treatment selection and enhancing the likelihood of beneficial outcomes [6]. Non-invasive brain signal acquisition techniques such as Electroencephalogram (EEG) can be used to extract these biomarkers. EEG is portable, has high temporal resolution, and is cost-effective making it an attractive tool for research. Machine learning algorithms and deep learning networks can integrate high-dimensional EEG data to predict treatment outcomes [7, 8]. In unipolar depression, baseline EEG power spectra in frontal channels predicted clinical response to tDCS [9].

In this study, we aimed to predict clinical remission to tDCS treatment in bipolar depression using pretreatment resting state eyes closed EEG signals. The power spectral density (PSD) of EEG signals, acquired before starting a home-based tDCS treatment course, were used to train deep learning models to predict remission.

## 2. Materials and Methods

### 2.1 Participant Recruitment and EEG Data Collection

All participants provided written informed consent for participation, the privacy rights of human participants have been observed, and the study was conducted in accordance with the World Medical Association Declaration of Helsinki. Ethical approval was provided by London - Fulham Research Ethics Committee on February 19^th^, 2022 (ref: 21/LO/0910). The study was an open-label, single arm trial of home-based tDCS treatment for bipolar depression [5]. Baseline EEG data had been acquired in a sub-sample of 21 participants with bipolar depression (14 women), mean age 51.38 ± 10.59 years. Diagnosis was made based on Diagnostic Statistical Manual of Mental Disorders, Fifth Edition (DSM- 5) criteria [1], determined by a structured assessment using the Mini-International Neuropsychiatric Interview (MINI; Version 7.0.2) [10]. Participants were in a current depressive episode of at least a moderate severity as defined by a minimum score of 18 on the Montgomery-Åsberg Depression Rating Scale (MADRS) (mean score 24.47±2.76). Participants were taking a stable dosage of mood stabilising medication for a minimum of two weeks or not taking any medications for a minimum of two weeks. Exclusion criteria included any concurrent psychiatric disorder, having a significant risk of suicide, or history of epilepsy. Full details of participation criteria and study details are reported with the trial outcomes [5].

tDCS treatment was 6-week home-based course of active tDCS, consisting of 5 sessions per week for the first 3 weeks followed by 2 sessions per week for 3 weeks, for a total of 21 sessions. Duration of each session was 30 minutes. tDCS was provided in bifrontal montage: anode at left dorsolateral prefrontal cortex (DLPFC) and cathode at right DLPFC (EEG electrode positions F3 and F4, respectively). Stimulation was 2 mA, and electrode area 23 centimetre square. During each session, the participant was seated comfortably with their eyes open, and a research team member was present by videoconference providing a discreet presence without interacting with the participant at each session

Treatment remission was defined as a MADRS score of 9 or less at the 6-week end of treatment study visit. Following tDCS treatment, 12 out of 21 participants achieved remission, while 9 participants did not meet remission criteria. The mean post-treatment MADRS score for the remission group was 5.08 ± 1.56, whereas the non-remission group had a mean score of 14.44 ± 4.87.

Two 5-minute pre-treatment, eyes-closed resting-state EEG recordings were acquired for each participant in their home environment using the Muse 2 EEG headband. This wireless system utilizes four dry electrodes positioned at AF7, AF8, TP9, and TP10, with FPz serving as the reference electrode. The EEG signals were sampled at 256 Hz. During the EEG recording, participants were instructed to sit relaxed with their eyes closed and without making any body movements. The recorded EEG file include timestamps for each sample and Horse Shoe Indicator (HSI) values indicate the quality of electrode connectivity. The HSI values 1, 2, and 4 signifies good, medium and poor connectivity respectively.

### 2.2. EEG Signal Pre-Processing and PSD extraction

Figure 1 illustrates the EEG signal pre-processing, PSD estimation and the subsequent deep learning-model training and leave-one-subject-out (LOSO) testing scheme. Each participant’s eyes-closed EEG signals were then divided into 60 non-overlapping windows, each lasting 10 seconds. To ensure good electrode connectivity and there by the quality of EEG, EEG windows with an average HSI below 2 were selected for PSD extraction. The PSD of the 10 seconds EEG windows were extracted using the Welch’s averaged modified periodogram method with smaller overlapping window of 3 seconds long and 1 second overlap. The 0 Hz frequency component in the PSD is constrained to zero to eliminate the impact of baseline shifts in the EEG signals.

**Figure 1.**
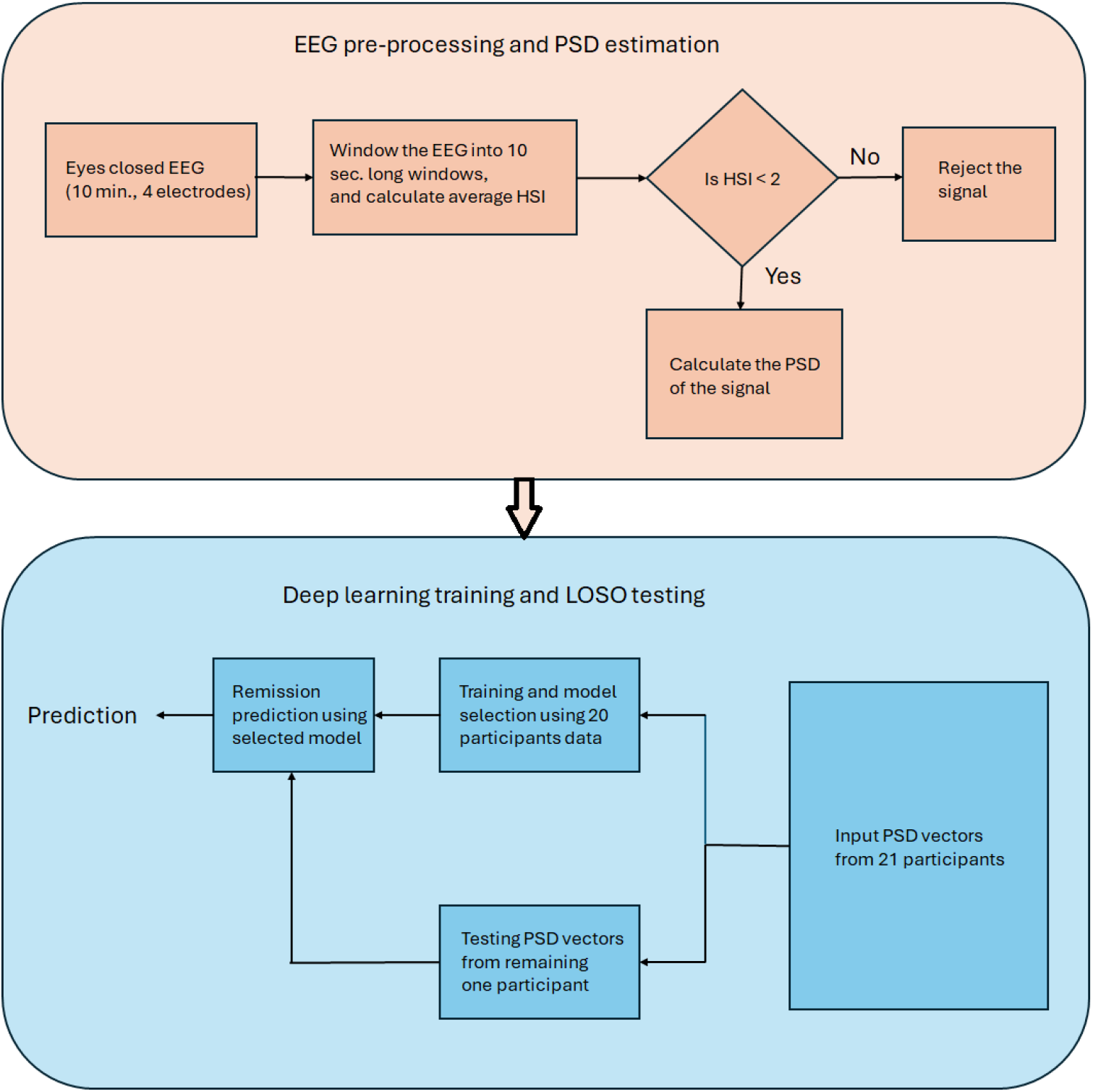
EEG pre-processing, PSD estimation and the deep learning-based treatment remission prediction scheme.

### 2.3. Deep learning model training and analysis

Deep learning models were developed to perform binary classification of generalized treatment remission versus non-remission using PSD derived from resting-state, eyes-closed EEG recordings. To evaluate the predictive differences in remission among PSD components across conventional EEG frequency bands, we individually used PSD vectors corresponding to the full band (0.5–60 Hz) as well as the delta (0.5–4 Hz), theta (4–8 Hz), alpha (8–12 Hz), beta (12–30 Hz), and gamma (30–60 Hz) bands as inputs to the deep learning architecture. Model performance was assessed for each PSD subset to determine the relative contribution of classical EEG bands to remission prediction. To further refine the evaluation, we examined combinations of EEG bands by concatenating PSD values from different bands, including all possible combinations of 2, 3, and 4 bands. Additionally, PSDs from individual electrodes and electrode combinations were tested as inputs to assess potential improvements in classification accuracy. Prior to model training and evaluation, each selected PSD input was standardized to have zero mean and unit variance.

The deep learning models examined are one-dimensional Convolutional Neural Networks (1DCNNs) [11], Long Short-Term Memory (LSTM) networks [12], Gated Recurrent Units (GRU) networks [13] and hybrid model combining CNN, LSTM/GRU, and multilayer perceptron. The architecture parameters like number of neurons or recurrent units in layers were selected accordance with the input dimension. Full band PSD extracted from EEG signals of four electrode has a dimension of 183 × 4. In this case, we utilized 61 filters with a kernel size of 3 for 1DCNN models. For individual bands, we employed 4 convolutional filters with a kernel size of 3 spanning full samples of input vector. For LSTM/GRU architectures with alpha band PSD with dimension 12 × 4, we employed 4 LSTM/GRU units for modelling the network. The PSD vectors from 4 electrodes inputted for EEG sub-bands has the following dimensions, delta band (12 × 4), theta band (12 × 4), alpha band (12 × 4), beta band (54 × 4), and gamma band (61 × 4).

For LSTM/GRU model, the first two layers used were LSTM/GRU layers. The final two layers of all the models were the same. The second to last layer was a fully connected dense layer with 64 neurons and the output layer was a single neuron output layer. For 1DCNN network, one convolution layer is used, for the hybrid models, one 1DCNN layer is used, and one LSTM/GRU layer is used prior to the last layers. Other than these layers, a maxpooling layer is used along with a 1DCNN layer, a flatten layer is used prior to the first dense layer, and dropout layers were used avoid overfitting. After evaluating treatment remission prediction using individualized EEG bands, we further assessed the performance of EEG band combinations. For this analysis, we employed the deep learning model that achieved the highest classification accuracy with individual EEG bands. The schematic diagram representing the hybrid model containing 1DCNN, GRU and dense layers are given in Figure 2.

**Figure 2.**
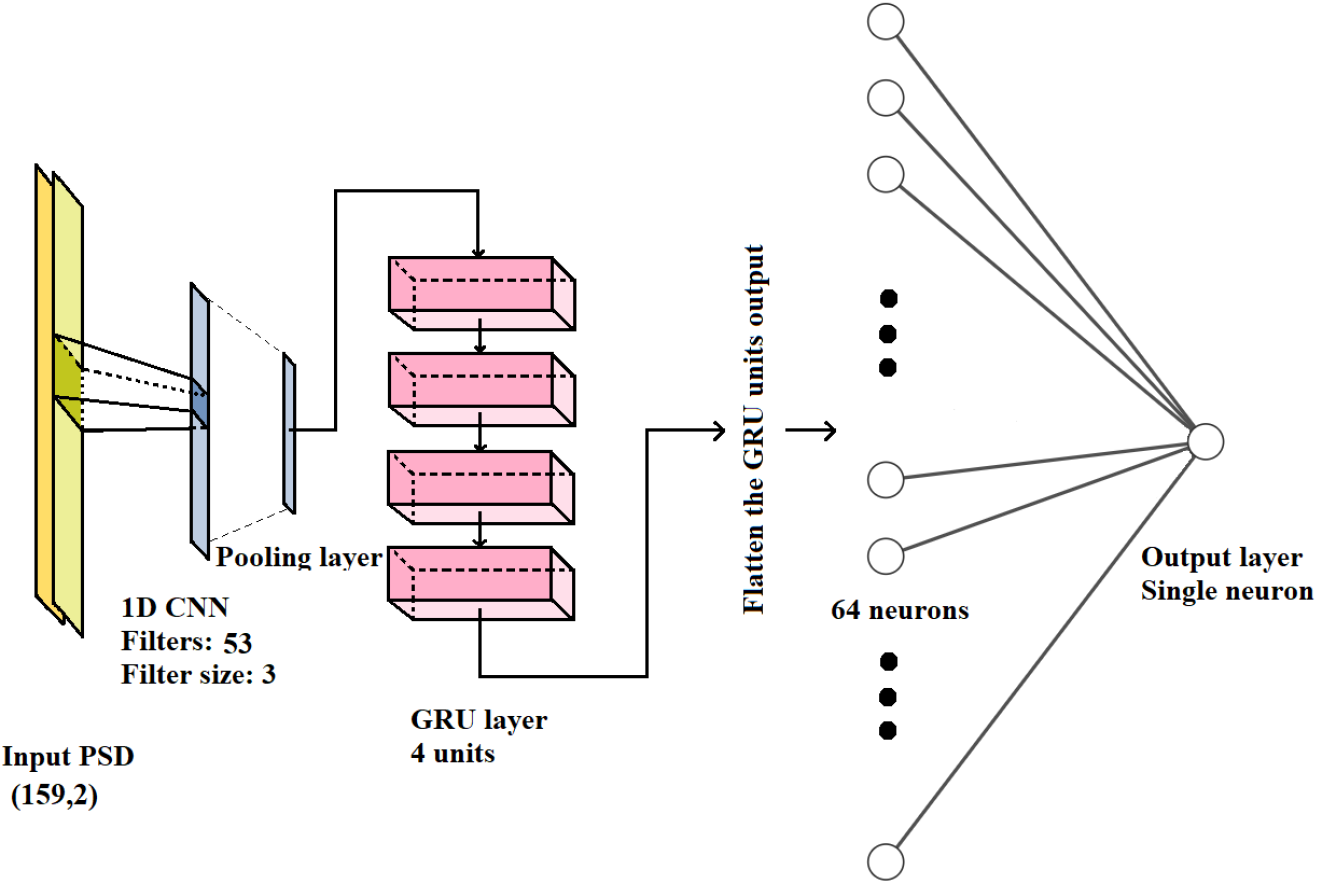
The hybrid model schematic containing 1DCNN, GRU and multilayer perceptron model.

Leave-one-subject-out (LOSO) testing method was used to report the performance matrices of the deep learning models. In each iteration, out of 21 participants, one participant was excluded for testing. From the remaining 20 participants, 4 (two from each class) were used for model selection/validation, and 16 for training. Deep learning model training was performed for 50 epochs, and the model with highest validation accuracy was selected for testing. This process was repeated for testing each participant, and the average testing accuracy obtained from 21 iterations were reported.

## 3. Results

Table 1 summarizes the averaged treatment remission prediction accuracies achieved and its standard deviation for 21 participants. The results obtained by using PSD inputs from different EEG frequency bands across various deep learning architectures were also specified in table 1. The PSD inputs from full band (0.3-60 Hz) gained superior classification accuracy compared to other EEG bands considered, the classification accuracies varied from 69-77% across different deep learning models. PSD inputs derived from the full frequency band (0.3–60 Hz) yielded superior classification accuracies relative to those obtained using other EEG sub-bands, with performance ranging from 69.59% to 76.43% across the evaluated deep learning models. For full band PSD input, the highest classification accuracy (76.43%) is obtained for hybrid model consisting of CNN-GRU architectures. Considering classification performance of individual classical EEG bands, gamma band yielded the highest classification accuracy (71%) using hybrid CNN-GRU model. The lowest classification accuracies were reported for delta band. It can also be observed from Table 1 that the standard deviation of the classification accuracies calculated among 21 participants are significantly high and is in a range between 17 to 30%.

**Table 1.** Prediction accuracies obtained for the different EEG frequency bands PSD values from four electrodes using different deep learning architectures.

| EEG frequency bands | Frequency range in Hz | Average prediction accuracy in % for different deep learning architectures |  |  |  |  |
| --- | --- | --- | --- | --- | --- | --- |
|  |  | CNN | LSTM | GRU | CNN & LSTM | CNN & GRU |
| Full band | 0.3-60 | 73.80 ±33.38 | 73.38±33.95 | 69.59±34.49 | 74.31±28.67 | 76.43±28.92 |
| Delta | 0.3-4 | 44.96±35.33 | 47.19±21.04 | 46.37±21.66 | 43.9±26.76 | 45.01±29.31 |
| Theta | 4-8 | 62.56±23.17 | 63.34±20.91 | 61.58±20.64 | 62.21±18.34 | 62.6±17.89 |
| Alpha | 8-12 | 53.49±31.23 | 54.33±22.59 | 57.2±21.62 | 51.23±24.18 | 53.01±31.75 |
| Beta | 12-30 | 63.36±35.68 | 60.83±31.33 | 61.63±30.6 | 64.06±27.25 | 67.32±26.45 |
| Gamma | 30-60 | 68.36±34.4 | 67.45±30.48 | 69.14±28.35 | 70.44±28.8 | 71±27.55 |

The classification accuracy has improved when PSD values from specific EEG bands were combined, and electrode selection performed. Table 2 shows the classification selected accuracies (accuracies >70%) for the combination of EEG band PSD values from AF7 and TP10 electrodes. The highest classification accuracy was obtained for the AF7, TP10 electrodes and for the combination of theta, beta and gamma band PSD input. The highest classification accuracy of 78.5% was obtained for the hybrid CNN, GRU model with sensitivity 81.4% and specificity 74.64%.

**Table 2.** Prediction accuracies obtained using CNN-GRU deep learning model for the selected (accuracy >70%) EEG frequency band PSD values from AF7, TP10 electrodes.

| EEG bands | Average prediction accuracy in % | Sensitivity in % | Specificity in % |
| --- | --- | --- | --- |
| Gamma | 74.83±28.97 | 79.11 | 69.13 |
| Delta, Gamma | 73.33±32.3 | 77.08 | 68.46 |
| Theta, Gamma | 76.8±27.55 | 78.69 | 74.27 |
| Alpha, Gamma | 70.15±31.96 | 76.18 | 62.11 |
| Beata, Gamma | 74.8±29.95 | 79.33 | 68.76 |
| Theta, Beta, Gamma | 78.5±28.2 | 81.4 | 74.64 |
| Alpha, Beta, Gamma | 75.46±26.85 | 76.86 | 73.59 |
| Delta, Alpha, Beta, Gamma | 74.83±26.64 | 74.57 | 75.19 |
| Theta, Alpha, Beta, Gamma | 71.92±30.09 | 76.77 | 65.44 |

## 4. Discussion

The PSD represents the power distribution of the signal in the frequency domain, with resolution limited by the sampling frequency. EEG signals are the result of potential difference arising from neuronal activity, within the microvolt range, typically encompass a frequency range of 0-60 Hz. In the context of EEG-based biomarkers for major depression and bipolar disorder, band power features and power-based asymmetry features are researched for diagnosing depression and forecasting treatment outcomes [14]. Although band power features are derived from the PSD, they represent only one way of representing PSD within a frequency range. By providing PSD as an input to deep learning architecture enables the model to extract features that can be localized within a limited frequency bandwidth or across multiple frequency bands.

Our results indicated that deep learning models, when applied to full-band PSD inputs, achieved higher classification accuracy compared to models using individual EEG bands. Specifically, hybrid models such as CNN-GRU and CNN-LSTM demonstrated notable performance, with accuracies of 73.25% and 76.43% respectively, indicating that the combination of PSDs different EEG bands will improve the treatment remission prediction. Among individual EEG bands, the gamma band yielded the highest classification accuracy. This may be due to the association of gamma oscillations in cognitive and emotional processing [15, 16].

Combining PSD values from multiple frequency bands for training offers advantages, such as learning features hidden in the relative changes of power distribution between multiple EEG bands. Additionally, a reduced input dimension compared to the full-band PSD helps avoid training problems like the curse of dimensionality. The highest accuracy was obtained when PSDs from theta, beta, and gamma bands of the AF7 and TP10 EEG electrodes were used. The prediction accuracy of 78.5% is obtained using the hybrid CNN-GRU model, with sensitivity (81.4%) and specificity (74.64%). The activity within the theta, beta, and gamma frequency bands has been implicated in various aspects of mood regulation, emotional processing, and mood disorders. In individuals with dysphoria, frontal theta activity is an EEG correlate of mood-related emotional processing [17] and bipolar patients exhibit a generalized increase in delta and theta activity [18]. Beta band activity is found to have correlation with severity of depression [19]. The delta and theta bands are associated with deeper stages of sleep, relaxation [20], blissful experiences during meditation [21], while the gamma band is often linked to higher-order cognitive functions [22]. Gamma oscillations have been found to distinguish between unipolar depression (major depressive disorder (MDD)) and healthy controls, as well as between bipolar disorder (BD) and MDD, and are altered by treatments [23]. Furthermore, repetitive transcranial magnetic stimulation (rTMS) treatment in MDD has been associated with significant increases in resting gamma power [24].

The results indicate that the combination of PSD inputs from the theta, beta and gamma bands from the AF7 and TP10 electrodes in hybrid deep learning models successfully identified biomarkers representing general brain arousal and specific cognitive processes relevant to mood regulation and treatment responsiveness.

The study’s limitations include the relatively small sample size (N=21) necessitates validation with larger cohorts to confirm the generalizability of our findings. Second, the use of a portable EEG system with only four dry electrodes (AF7, AF8, TP9, TP10) presents technical constraints: (1) reduced spatial resolution compared to high-density arrays, (2) increased susceptibility to ocular artifacts in frontal regions, and (3) potentially lower signal quality than conventional wet electrode systems. Employing clinical-grade, high-density wet electrode systems, incorporating multimodal clinical features may enhance predictive accuracy. These limitations highlight important trade-offs between home-based recording and signal quality that should inform future translational applications.

## 6. Conclusion

In conclusion, our study demonstrated the potential of using deep learning models with power spectral density extracted from baseline resting-state EEG to predict treatment remission of home-based tDCS treatment in bipolar depression. Home-based tDCS treatment combined with advanced analytical methods could provide the opportunity for personalised treatment decision-making approaches. Validation with larger datasets is required to establish the reliability and generalizability in real-world clinical applications.

## Data Availability

All data produced in the present study are available upon reasonable request to the authors

